# Monitoring Report: GLP-1 RA Prescribing Trends - September 2026 Data

**DOI:** 10.1101/2025.03.06.25323524

**Authors:** Samuel Gratzl, Karen Gilbert Farrar, Emma Holler, Madhura S Vachon, Nina Masters, Zachary Wallen, Brianna M Goodwin Cartwright

## Abstract

**Background:** Limited recent data exist on prescribing patterns and patient characteristics for glucagon-like peptide 1 receptor agonists (GLP-1 RAs), an important drug class used as anti-diabetic medication (ADM) for patients with type 2 diabetes mellitus (T2D) and/or anti-obesity medication (AOM) in patients with overweight or obesity.

For brevity, we use the term GLP-1 RA to refer to both GLP-1 RA and dual GLP-1 RA/GIP medications.

**Objective:** To describe recent trends in prescribing and dispensing of GLP-1-based medications in the US.

**Methods:** Using a subset of real-world electronic health record (EHR) data from Truveta, a growing collective of health systems that provide more than 18% of all daily clinical care in the US, we identified people who were prescribed a GLP-1-based medication between January 01, 2019 and September 30, 2026. We describe prescribing volumes and patient characteristics over time, by medication, and by FDA-labeled use. Among the subset of patients for whom post-prescription dispensing data is available, we describe the proportion and characteristics of patients who were and were not dispensed a GLP-1 RA following their prescription.

**Results:** 4,458,532 patients were prescribed a GLP-1 RA between January 2019 and September 2026, with 23,616,330 total prescriptions during this period. Overall prescribing rates (GLP-1 RA prescriptions as a proportion of all prescriptions) increased by 7.0% from June to September 2026. AOM prescribing increased by 12.2% (+0.38 percentage points), with the largest month-over-month increase occurring from June to July and prescribing remained relatively steady thereafter. On July 1, 2026, the Medicare GLP-1 Bridge Program was implemented to expand access to GLP-1 AOMs for Medicare beneficiaries by reducing out-of-pocket costs. The June-to-July increase coincided with the July 1 implementation of the Medicare GLP-1 Bridge and may reflect expanded access to GLP-1 RAs for weight management through the program. This increase was largely driven by AOM tirzepatide, for which prescribing increased by 16.9%. Although AOM orforglipron prescribing more than doubled (+112.2%), orforglipron still accounted for a small proportion of AOM prescriptions. Tirzepatide remained the most commonly prescribed GLP-1 RA, while semaglutide prescribing remained relatively stable. Among first-time prescriptions for which indication could be established, ADMs accounted for 62.6% and AOMs accounted for 37.4%. Overall, first-time prescribing increased from June to July before declining through August and September, with rates in September similar to June (-1.8%). First-time AOM prescribing also increased from June to July before declining, with rates in September remaining relatively stable compared with June (+3.7%). First-time AOM tirzepatide prescribing increased by 10.1% from June to September, while first-time AOM semaglutide prescribing decreased by 12.5%.

**Trends in Prescribing:** 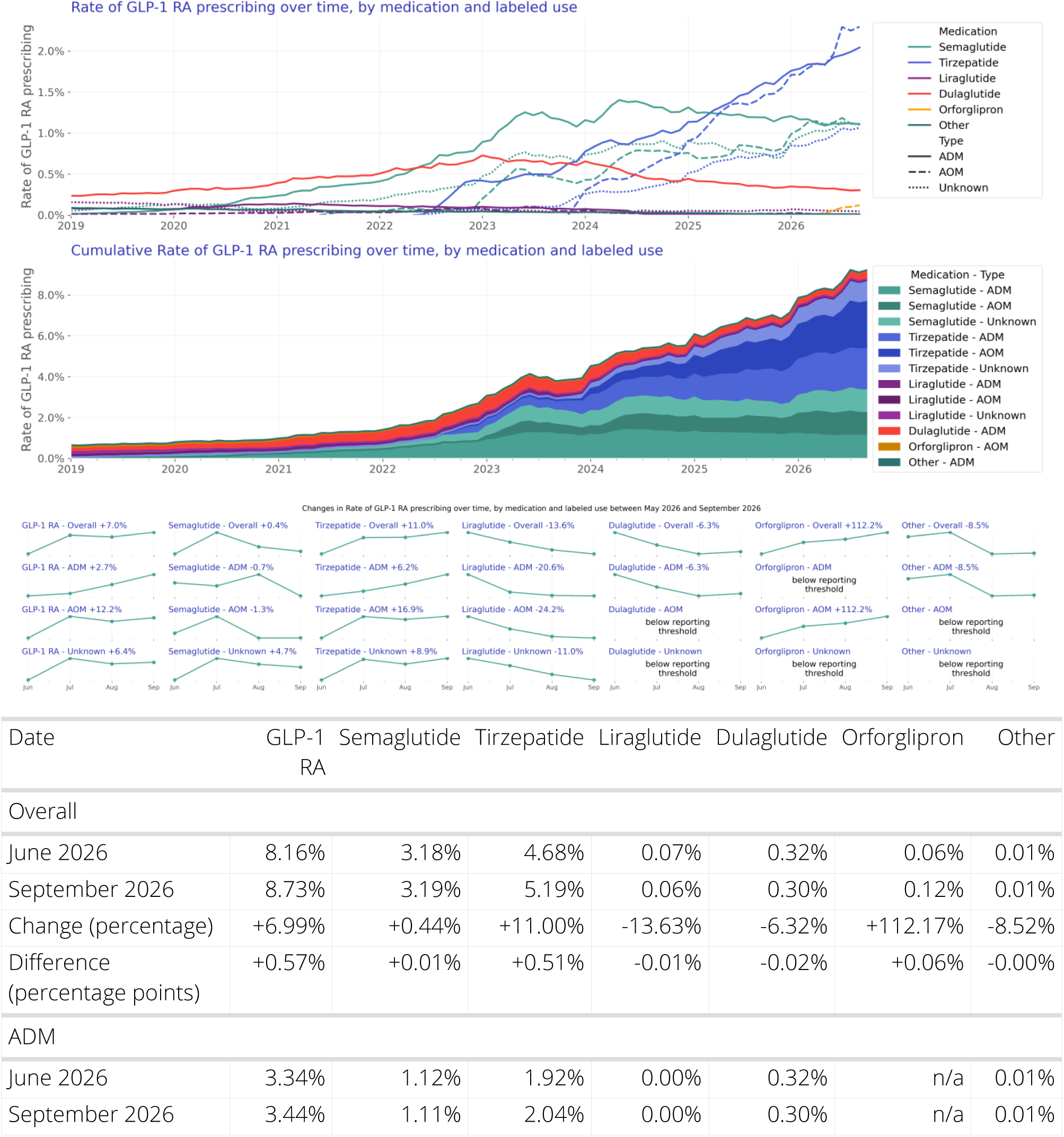

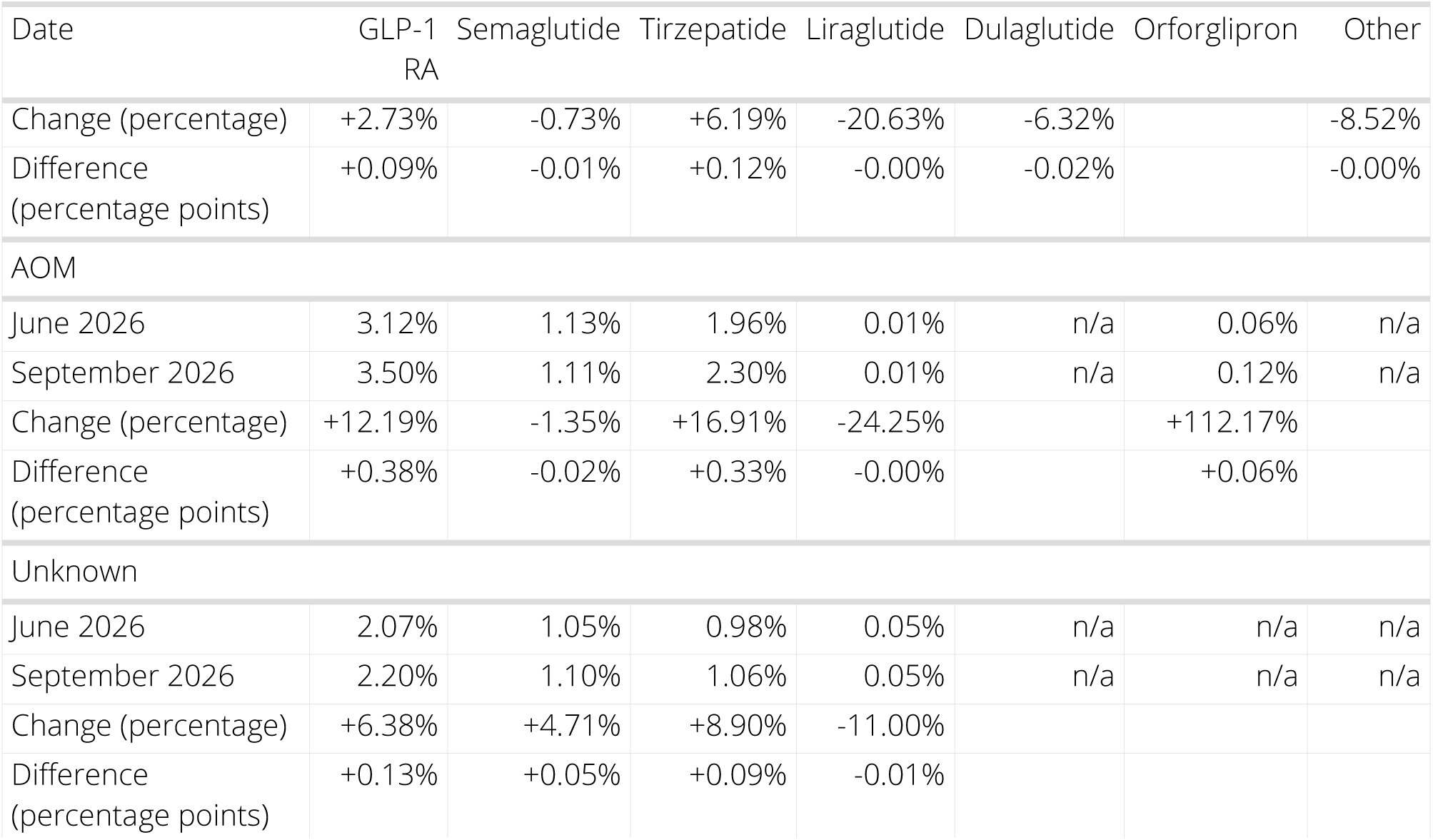

- Overall prescribing rates (GLP-1 RA prescriptions per total prescriptions) in September 2026 increased slightly relative to June 2026 (+6.99%).
- ADM prescribing showed slight month-to-month increases, however ADM prescribing in September 2026 are about the same June 2026 (+2.73%).
- Month-over-month AOM prescribing increased most from June to July 2026 and remained relatively steady through September 2026. Overall, there was a 0.38 percentage point increase compared with June 2026 (+12.2%). The largest increase came from AOM tirzepatide prescriptions (+16.9%), which accounted for most of the overall increase. AOM orforglipron prescribing more than doubled (+112.2%), although overall prescribing remained low at 0.12% in September 2026.
- The June-to-July increase in AOM prescribing coincided with the July 1 implementation of the Medicare GLP-1 Bridge and may reflect expanded access to GLP-1 RAs for weight management through the program.
- Tirzepatide remains the most commonly prescribed GLP-1 RA and continued to increase over the last quarter, particularly when prescribed for obesity, while semaglutide prescribing remained relatively stable.

**Trends in First-Time Prescribing:** 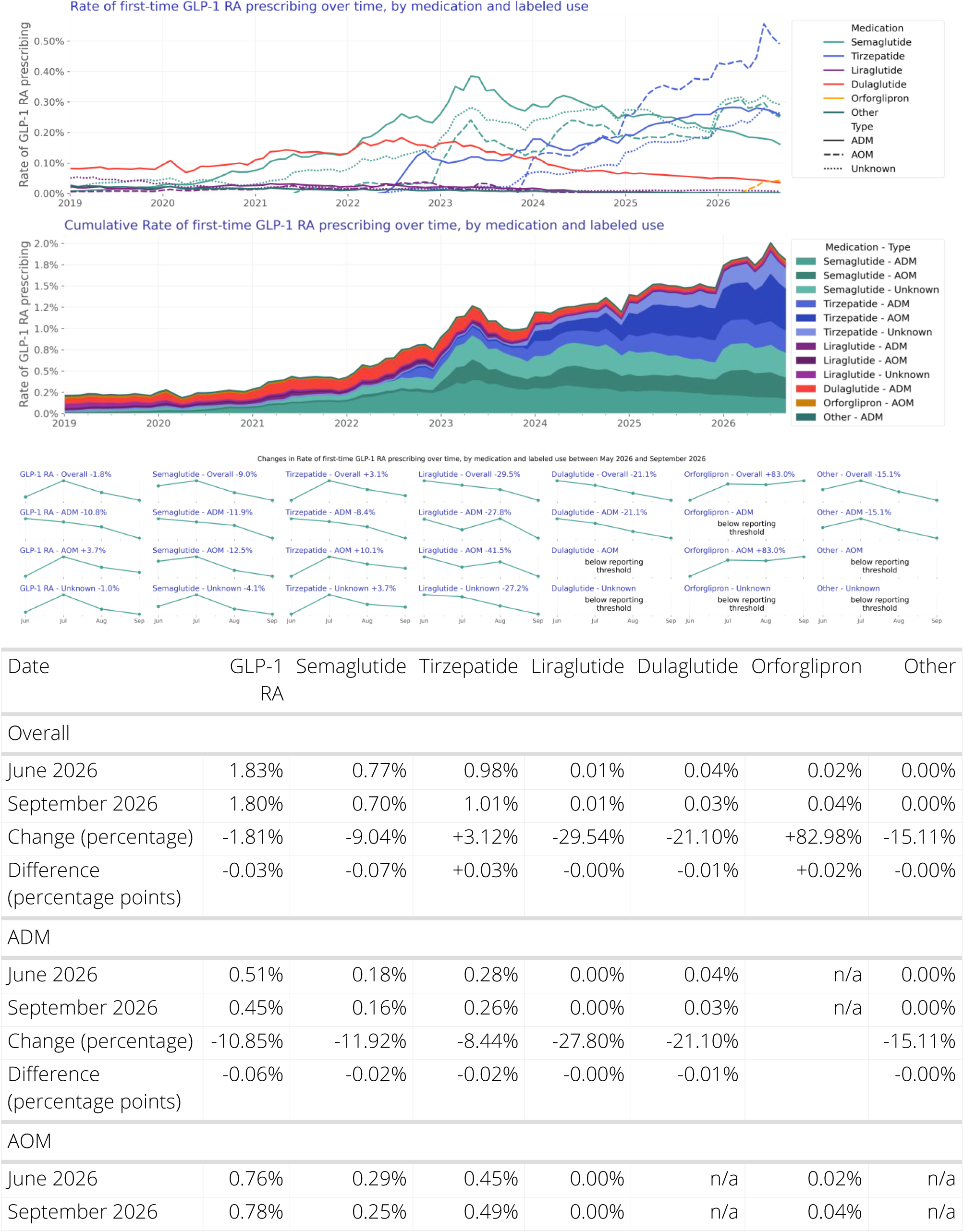

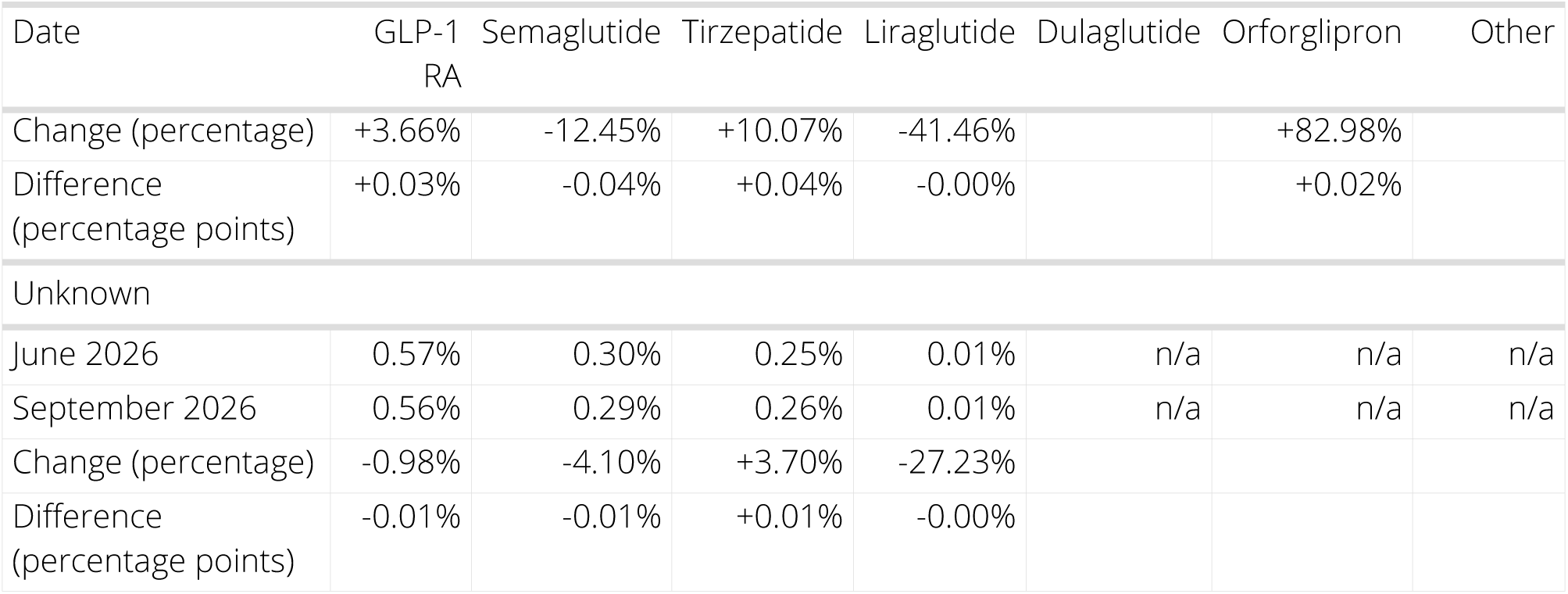

- Overall first-time GLP-1 RA prescribing increased from June to July 2026, then declined in August and September. By September, first-time prescribing rates were similar to June (-1.81%).
- First-time prescribing of ADMs decreased in September 2026, relative to June 2026 (-10.85%).
- First-time AOM prescribing increased from June to July 2026, then declined and remained relatively steady through September. Overall, first-time AOM prescribing in September was similar to June (+3.66%). First-time AOM tirzepatide prescribing increased 10.1%, while AOM orforglipron prescribing nearly doubled (+83.0%), although overall first-time prescribing of AOM orforglipron remained low.
- Similar to overall prescribing, the June-to-July increase in first-time AOM prescribing coincided with the July 1 implementation of the Medicare GLP-1 Bridge and may reflect expanded access to GLP-1 RAs for weight management through the program.

**Trends in Dispensing:** 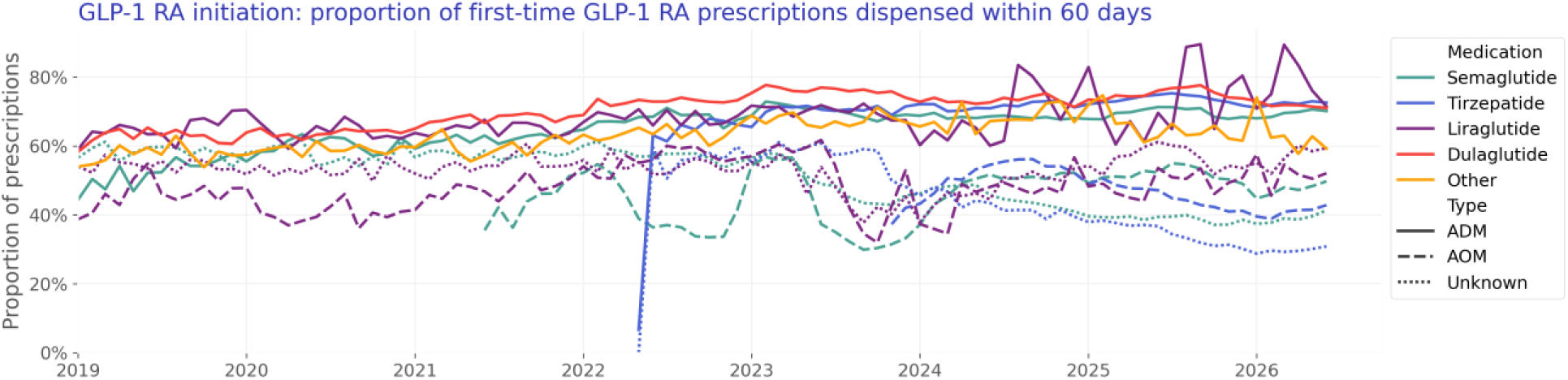

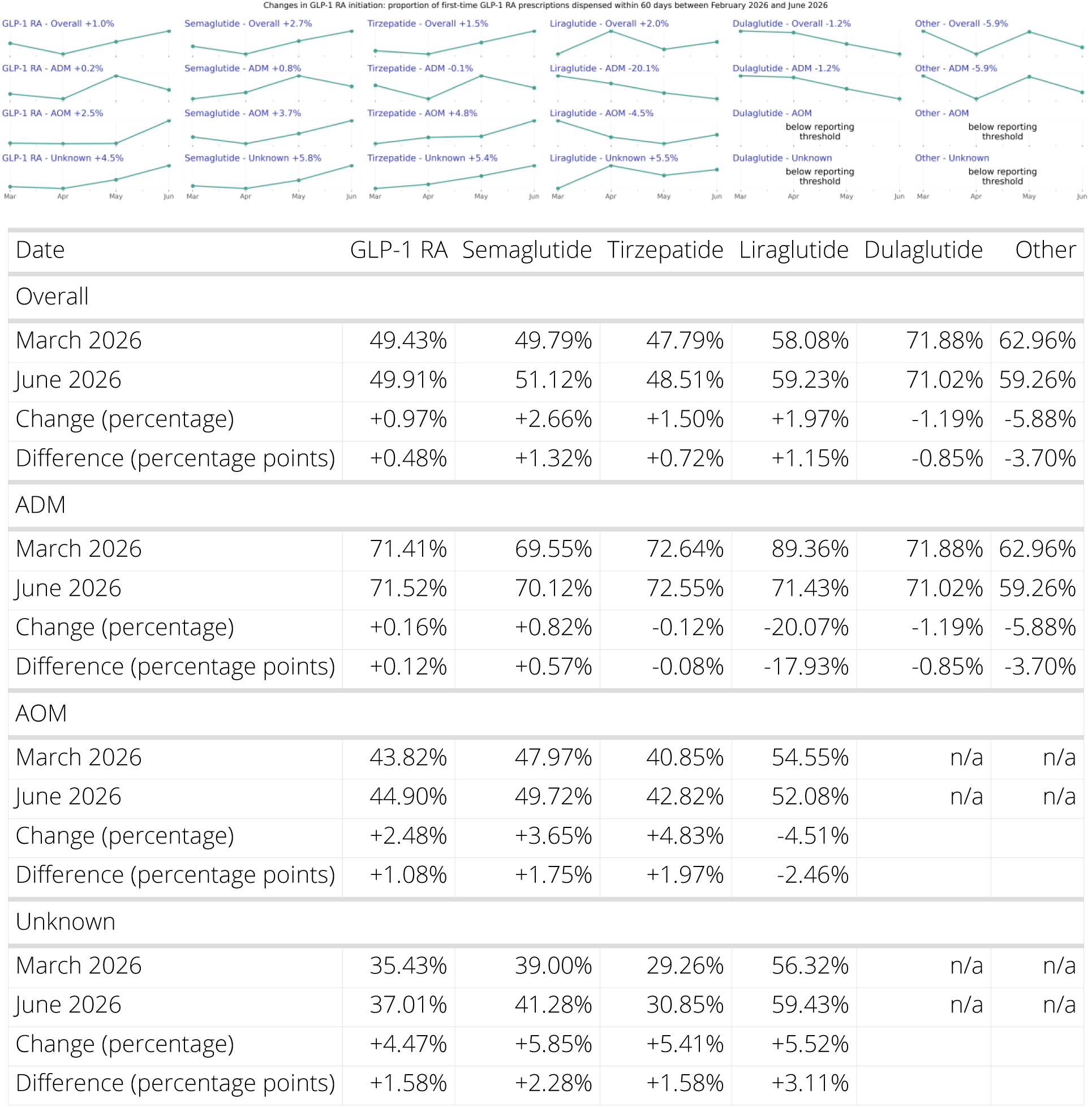

- During the full time period, 69.9% of GLP-1 RA prescriptions had a dispense within 60 days of their prescription (e.g., were initiated within 60 days).
- For patients first prescribed an ADM in June 2026, 71.5% filled a GLP-1 RA within 60 days. Initiation rates for ADM GLP-1 RA are about the same in June 2026, relative to March 2026 (+0.16%).
- For patients first prescribed an AOM in June 2026, 44.9% filled a GLP-1 RA within 60 days. Initiation rates for AOM GLP-1 RA are about the same in June 2026, relative to March 2026 (+2.48%).

**About this report:** This report provides recent trends in the prescribing and dispensing of glucagon-like peptide-1 receptor agonist (GLP-1 RAs)-based medications, including dual GLP-1 RA/gastric inhibitory polypeptide (GIP) over time, by medication, and by FDA-labeled use. We used near real-time, aggregated electronic health record (EHR) data from a collective of US healthcare systems to describe prescribing and dispensing of GLP-1 RA medications from January 2019 to September 2026.

## Introduction

Obesity and type 2 diabetes mellitus (T2D) are interconnected epidemics with a high prevalence and individual and combined negative impacts on health (CDC, 2020, Hales, 2020). Glucagon-like peptide-1 receptor agonist (GLP-1 RAs) -based medications, including dual GLP-1 RA/gastric inhibitory polypeptide (GIP), have been shown to improve glycemic control in patients with T2D and reduce weight in patients with and without T2D (Davies, 2015, Davies, 2021, Garvey, 2023, Jastreboff, 2022, Wilding, 2023, Yeh, 2023). GLP-1 RA have also demonstrated cardiovascular benefit in patients with and without T2D (Lincoff, 2023, Ussher, 2023).

Interest in these medications has recently accelerated, largely for their weight-loss effects; however, limited empiric data exist on prescribing, dispensing, and patient characteristics. Access to and use of GLP-1 RA may be impacted by high cost, limited insurance coverage for patients without T2D and medication shortages.

## Methods

### Data

We evaluated prescribing and dispensing of GLP-1 RAs between January 2019 and September 2026 using a subset of Truveta Data. Truveta provides access to continuously updated, linked, and de-identified electronic health record (EHR) from a collective of US health care systems that provide 18% of all daily clinical care in the US, including structured information on demographics, encounters, diagnoses, vital signs (e.g., weight, BMI, blood pressure), medication requests (prescriptions), medication administration, laboratory and diagnostic tests and results (e.g., HbA1c tests and values), and procedures. Updated EHR data are provided daily to Truveta by member health care systems. In addition to EHR data for care delivered within Truveta member health care systems, medication dispensing and social drivers of health (SDOH) information are made available through linked third-party data. Medication dispense (via e-prescribing data) includes fills for prescriptions written both within and outside Truveta member health care systems, providing greater observability into patients’ medication history. Medication dispense histories are updated at the time of the encounter and include fill dates, NDC or RxNorm codes, quantity dispensed, and days of medication supplied.

### Population

We identified GLP-1 RA prescriptions (medication requests) and dispenses among adults using RxNorm and NDC codes for dulaglutide, exenatide, liraglutide, lixisenatide, semaglutide, and tirzepatide. Exenatide and lixisenatide were grouped as ‘other’ given small relative sizes. Brands were identified when possible using RxNorm and NDC codes. Brands labeled for treatment of T2D were classified as ADMs and brands labeled for treatment of overweight and obesity were classified as AOMs. When brand could not be established, the labeled use was classified as unknown.

### Prescribing Analysis

Characteristics of patients ever prescribed a GLP-1 RA were summarized, including demographics, social drivers of health (SDOH), and presence of T2D and/or overweight or obesity. Characteristics are provided by medication and by year of first prescription.

Overall and first-time prescribing rates of GLP-1 RA were summarized over time by medication and labeled use. The prescribing rate was calculated monthly as the number of patients with a GLP-1 RA prescription divided by the number of patients with any prescription in the month. Absolute differences and percentage changes in the rate of GLP-1 RA prescribing over the most recent 3-month period were summarized overall and by medication and labeled use.

### Dispensing Analysis

We describe the proportion of GLP-1 RA prescriptions (overall and first-time) with a dispense occurring within 60 days. Dispensing analyses were restricted to prescriptions for which complete 60-day dispensing outcomes could be observed. Dispensing outcomes were considered observable if (1) a dispense was recorded in the 60 days following the prescription, and/or (2) an encounter took place after the 60-day period ended (at which time any dispenses would be observed). To allow sufficient follow-up, we report dispensing with a 90-day delay. Given shortages and expected substitutions at the pharmacy, we considered the dispense of any GLP-1 RA, which could differ from the prescribed GLP-1 RA.

We plot the proportion of prescriptions (overall and first-time) dispensed within 60 days over time, by medication and use. Differences in dispensing were expected between ADMs and AOMs given ADMs are typically covered by insurance, whereas insurance coverage for AOMs is limited. As such, we provide dispensing rates stratified by FDA labeled use. Further, we stratify dispensing by age 65+ vs. 18-64 as a proxy for Medicare coverage. Medicare never provides coverage for AOMs, whereas commercial insurers may.

## Results

### Overall population

The demographics of patients by first-prescribed medication are as follows:

**Table 1:** Patient Characteristics by First-Prescribed GLP-1 RA Medication.

|  | Semaglutide | Tirzepatide | Liraglutide | Dulaglutide | Orforglipron | Other | Overall |
| --- | --- | --- | --- | --- | --- | --- | --- |
| N | 2,237,185<br>(100.00%) | 1,412,552<br>(100.00%) | 182,296<br>(100.00%) | 451,266<br>(100.00%) | 8,795<br>(100.00%) | 42,675<br>(100.00%) | 4,334,769<br>(100.00%) |
| Age Group |  |  |  |  |  |  |  |
| 18-44 | 595,862<br>(26.63%) | 427,065<br>(30.23%) | 52,842<br>(28.99%) | 62,112<br>(13.76%) | 2,072<br>(23.56%) | 4,401<br>(10.31%) | 1,144,354<br>(26.40%) |
| 45-64 | 1,048,778<br>(46.88%) | 665,591<br>(47.12%) | 89,305<br>(48.99%) | 222,500<br>(49.31%) | 3,078<br>(35.00%) | 18,700<br>(43.82%) | 2,047,952<br>(47.24%) |
| 65+ | 592,545<br>(26.49%) | 319,896<br>(22.65%) | 40,149<br>(22.02%) | 166,654<br>(36.93%) | 3,645<br>(41.44%) | 19,574<br>(45.87%) | 1,142,463<br>(26.36%) |
| Sex |  |  |  |  |  |  |  |
| Female | 1,465,670<br>(65.51%) | 938,858<br>(66.47%) | 127,625<br>(70.01%) | 244,150<br>(54.10%) | 6,360<br>(72.31%) | 22,350<br>(52.37%) | 2,805,013<br>(64.71%) |
| Male | 771,515<br>(34.49%) | 473,694<br>(33.53%) | 54,671<br>(29.99%) | 207,116<br>(45.90%) | 2,435<br>(27.69%) | 20,325<br>(47.63%) | 1,529,756<br>(35.29%) |
| Race |  |  |  |  |  |  |  |
| White | 1,601,993<br>(71.61%) | 1,048,435<br>(74.22%) | 130,128<br>(71.38%) | 302,334<br>(67.00%) | 6,870<br>(78.11%) | 30,771<br>(72.11%) | 3,120,531<br>(71.99%) |
| Black or African American | 327,100<br>(14.62%) | 169,559<br>(12.00%) | 27,790<br>(15.24%) | 78,355<br>(17.36%) | 734 (8.35%) | 5,740<br>(13.45%) | 609,278<br>(14.06%) |
| Asian | 54,395<br>(2.43%) | 30,042<br>(2.13%) | 2,984<br>(1.64%) | 11,859<br>(2.63%) | 194 (2.21%) | 955<br>(2.24%) | 100,429<br>(2.32%) |
| Unknown | 153,053<br>(6.84%) | 106,458<br>(7.54%) | 11,859<br>(6.51%) | 35,328<br>(7.83%) | 654 (7.44%) | 3,118<br>(7.31%) | 310,470<br>(7.16%) |
| Native Hawaiian or Other Pacific Islander | 7,177<br>(0.32%) | 3,932<br>(0.28%) | 713<br>(0.39%) | 1,795<br>(0.40%) | 16 (0.18%) | 156<br>(0.37%) | 13,789<br>(0.32%) |
| American Indian or Alaska Native | 16,130<br>(0.72%) | 7,594<br>(0.54%) | 2,246<br>(1.23%) | 3,366<br>(0.75%) | 36 (0.41%) | 392<br>(0.92%) | 29,764<br>(0.69%) |
| Other Race | 77,337<br>(3.46%) | 46,532<br>(3.29%) | 6,576<br>(3.61%) | 18,229<br>(4.04%) | 291 (3.31%) | 1,543<br>(3.62%) | 150,508<br>(3.47%) |
| Ethnicity |  |  |  |  |  |  |  |
| Hispanic or Latino | 234,300<br>(10.47%) | 142,481<br>(10.09%) | 18,833<br>(10.33%) | 55,358<br>(12.27%) | 656 (7.46%) | 4,799<br>(11.25%) | 456,427<br>(10.53%) |
| Multiethnic | 35 (0.00%) | 21 (0.00%) | n/a | 8 (0.00%) | n/a | 2 (0.00%) | 66<br>(0.00%) |
| Not Hispanic or Latino | 1,690,250<br>(75.55%) | 1,069,496<br>(75.71%) | 138,685<br>(76.08%) | 338,619<br>(75.04%) | 6,824<br>(77.59%) | 32,297<br>(75.68%) | 3,276,171<br>(75.58%) |
| Unknown | 312,600<br>(13.97%) | 200,554<br>(14.20%) | 24,778<br>(13.59%) | 57,281<br>(12.69%) | 1,315<br>(14.95%) | 5,577<br>(13.07%) | 602,105<br>(13.89%) |
| Characteristics |  |  |  |  |  |  |  |
| Type II Diabetes | 864,843<br>(38.66%) | 392,203<br>(27.77%) | 75,168<br>(41.23%) | 315,215<br>(69.85%) | 686 (7.80%) | 30,261<br>(70.91%) | 1,678,376<br>(38.72%) |
| Overweight or Obesity | 1,546,362<br>(69.12%) | 1,002,073<br>(70.94%) | 114,782<br>(62.96%) | 236,103<br>(52.32%) | 7,678<br>(87.30%) | 20,329<br>(47.64%) | 2,927,327<br>(67.53%) |
| Labeled Use |  |  |  |  |  |  |  |
| ADM | 895,116<br>(40.01%) | 523,558<br>(37.06%) | 62,742<br>(34.42%) | 451,266<br>(100.00%) | n/a | 42,675<br>(100.00%) | 1,975,357<br>(45.57%) |
| AOM | 547,288<br>(24.46%) | 573,741<br>(40.62%) | 50,121<br>(27.49%) | n/a | 8,795<br>(100.00%) | n/a | 1,179,945<br>(27.22%) |
| Unknown | 794,781<br>(35.53%) | 315,253<br>(22.32%) | 69,433<br>(38.09%) | n/a | n/a | n/a | 1,179,467<br>(27.21%) |
| Year |  |  |  |  |  |  |  |
| 2019 | 18,976<br>(0.85%) | n/a | 26,594<br>(14.59%) | 31,622<br>(7.01%) | n/a | 8,333<br>(19.53%) | 85,525<br>(1.97%) |
| 2020 | 35,135<br>(1.57%) | n/a | 23,391<br>(12.83%) | 38,081<br>(8.44%) | n/a | 7,074<br>(16.58%) | 103,681<br>(2.39%) |
| 2021 | 91,454<br>(4.09%) | n/a | 31,589<br>(17.33%) | 65,368<br>(14.49%) | n/a | 6,782<br>(15.89%) | 195,193<br>(4.50%) |
| 2022 | 216,942<br>(9.70%) | 33,466<br>(2.37%) | 39,095<br>(21.45%) | 96,415<br>(21.37%) | n/a | 6,837<br>(16.02%) | 392,755<br>(9.06%) |
| 2023 | 488,391<br>(21.83%) | 97,256<br>(6.89%) | 32,479<br>(17.82%) | 96,927<br>(21.48%) | n/a | 5,846<br>(13.70%) | 720,899<br>(16.63%) |
| 2024 | 520,594<br>(23.27%) | 269,038<br>(19.05%) | 13,402<br>(7.35%) | 58,168<br>(12.89%) | n/a | 4,266<br>(10.00%) | 865,468<br>(19.97%) |
| 2025 | 469,988<br>(21.01%) | 513,881<br>(36.38%) | 9,438<br>(5.18%) | 41,614<br>(9.22%) | n/a | 2,478<br>(5.81%) | 1,037,399<br>(23.93%) |
| 2026 | 395,705<br>(17.69%) | 498,911<br>(35.32%) | 6,308<br>(3.46%) | 23,071<br>(5.11%) | 8,795<br>(100.00%) | 1,059<br>(2.48%) | 933,849<br>(21.54%) |
| Comorbidity |  |  |  |  |  |  |  |
| Asthma | 309,962<br>(13.86%) | 203,272<br>(14.39%) | 26,437<br>(14.50%) | 50,564<br>(11.20%) | 1,413<br>(16.07%) | 4,025<br>(9.43%) | 595,673<br>(13.74%) |
| CKD | 290,030<br>(12.96%) | 134,323<br>(9.51%) | 26,363<br>(14.46%) | 88,118<br>(19.53%) | 1,068<br>(12.14%) | 8,517<br>(19.96%) | 548,419<br>(12.65%) |
| COPD | 114,300<br>(5.11%) | 60,887<br>(4.31%) | 9,975<br>(5.47%) | 32,367<br>(7.17%) | 473 (5.38%) | 3,081<br>(7.22%) | 221,083<br>(5.10%) |
| Hyperlipidemia | 1,119,871<br>(50.06%) | 669,378<br>(47.39%) | 75,029<br>(41.16%) | 237,239<br>(52.57%) | 5,423<br>(61.66%) | 22,005<br>(51.56%) | 2,128,945<br>(49.11%) |
| Hypertension | 1,152,600<br>(51.52%) | 666,170<br>(47.16%) | 87,541<br>(48.02%) | 260,793<br>(57.79%) | 4,605<br>(52.36%) | 24,775<br>(58.06%) | 2,196,484<br>(50.67%) |
| Major Depression | 261,563<br>(11.69%) | 159,977<br>(11.33%) | 23,434<br>(12.85%) | 40,486<br>(8.97%) | 1,186<br>(13.48%) | 3,370<br>(7.90%) | 490,016<br>(11.30%) |

### Prescribing over time

#### Overall

We describe the monthly rate of prescriptions by medication and FDA labeled use over time:

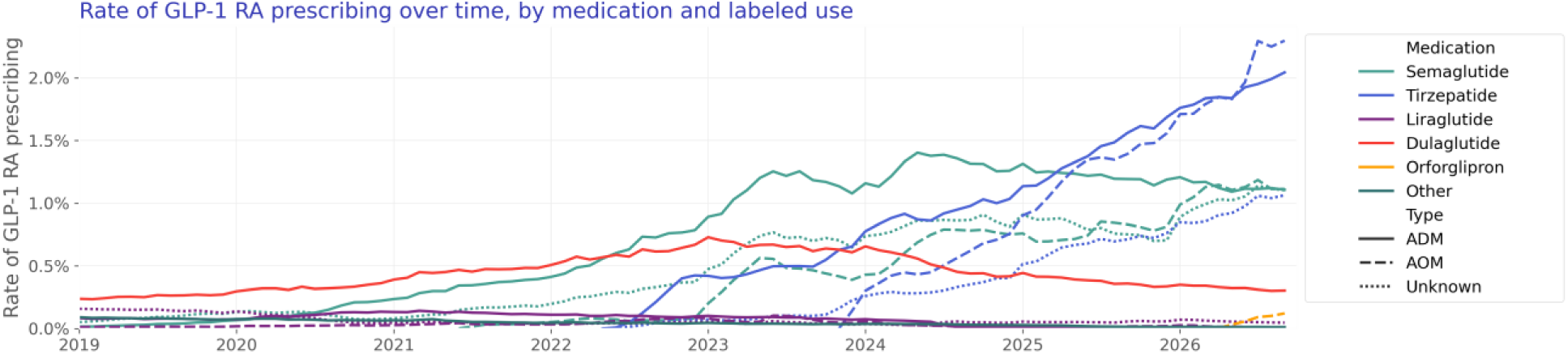
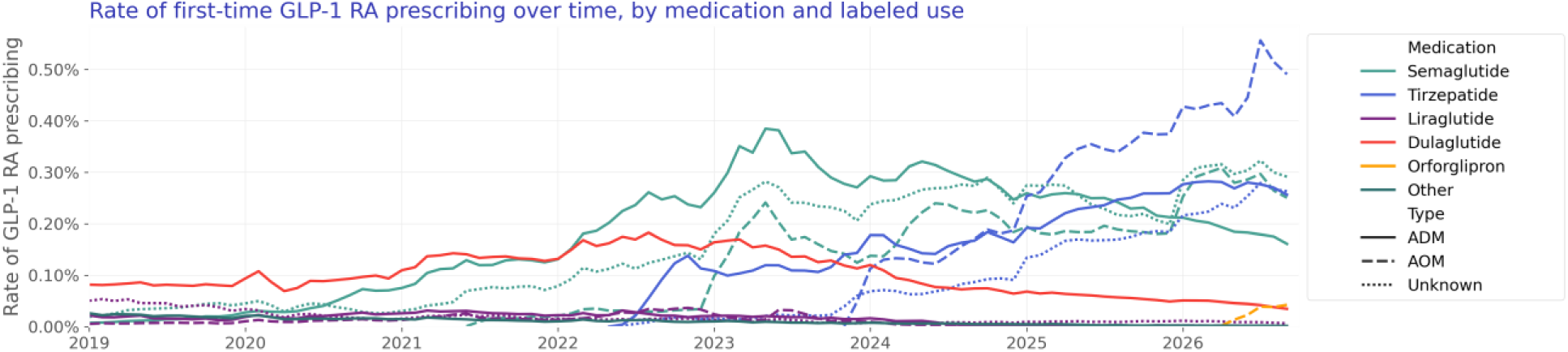

#### First-time

We describe the monthly rate of first-time prescriptions by medication and FDA labeled use over time.

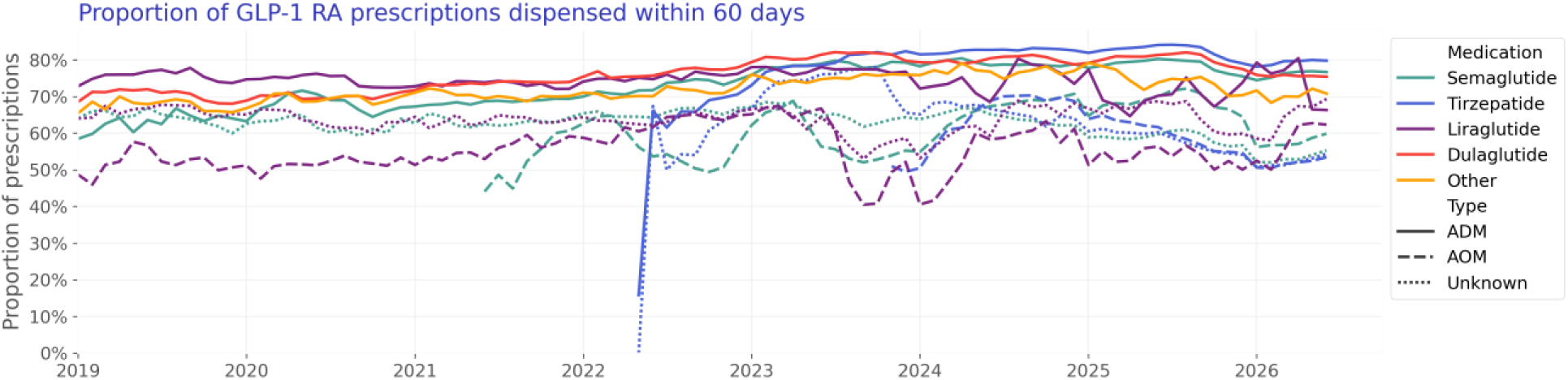

Characteristics of patients by year of first GLP-1 RA use are as follows:

**Table 2:**
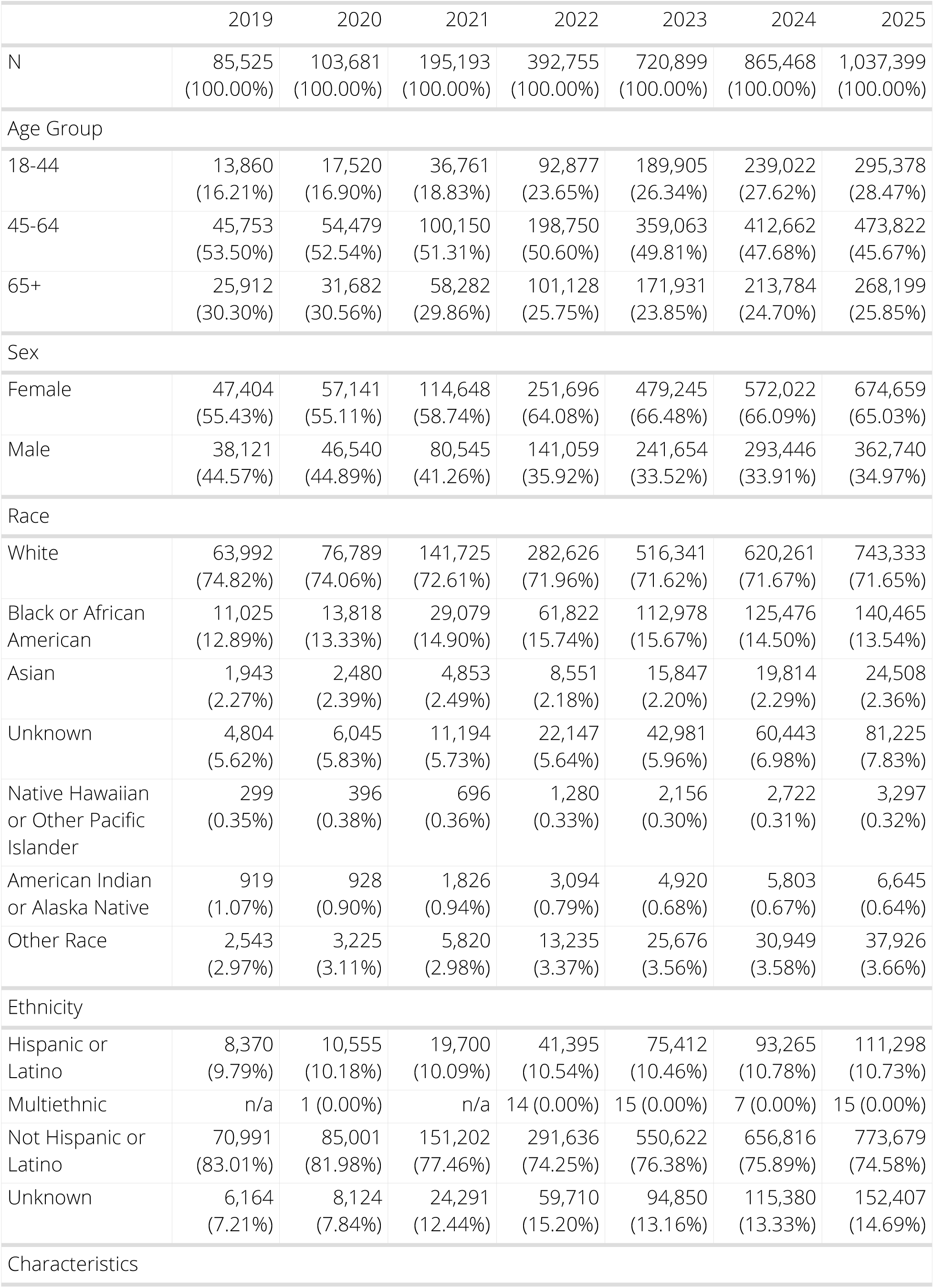
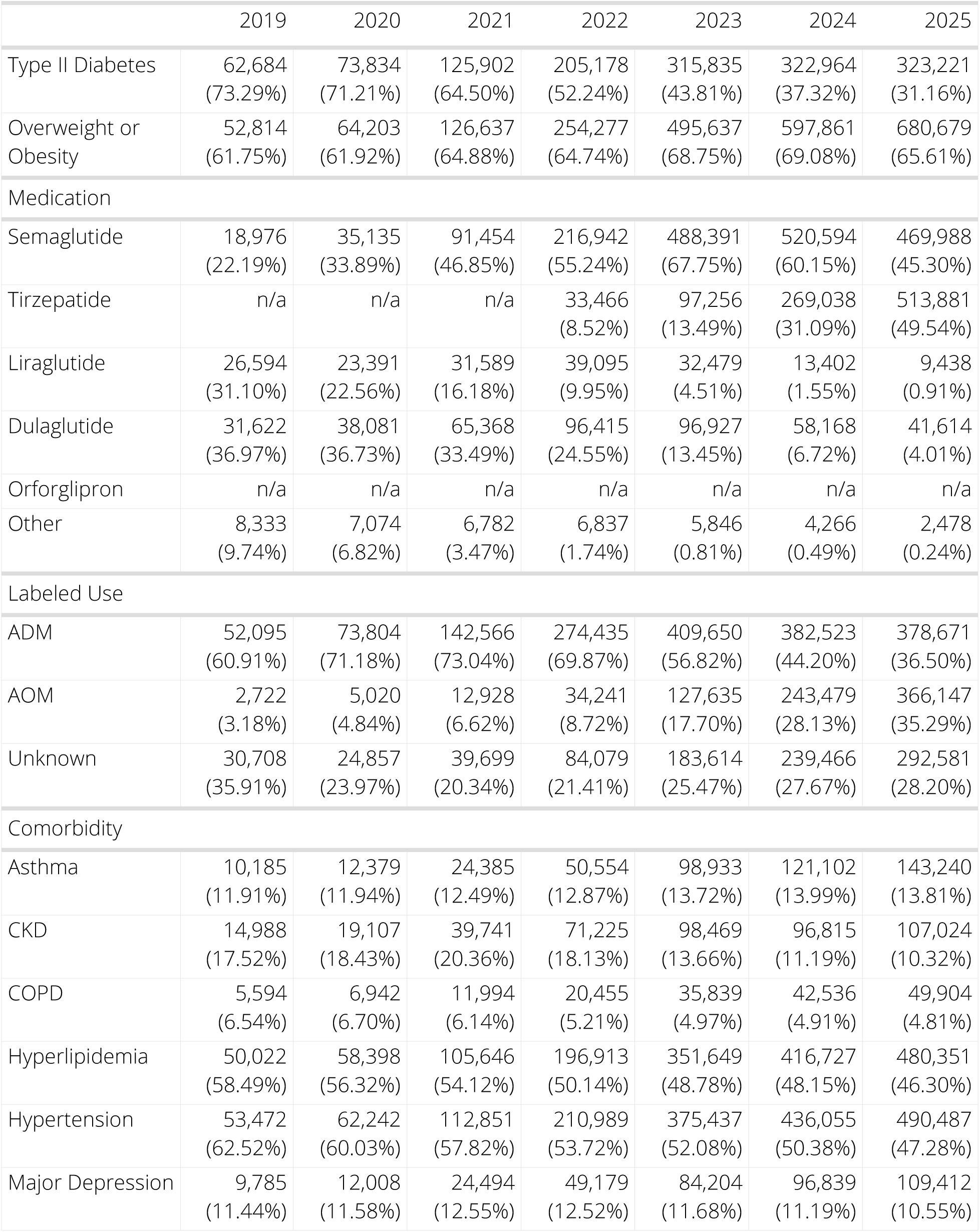
Patient Characteristics by Year of First GLP-1 RA Prescription.

### 60-day dispensing over time

We describe the proportion of patients with any GLP-1 RA prescriptions (first-time and repeat) in a given month who filled any GLP-1 RA in the 60 days following their prescription.

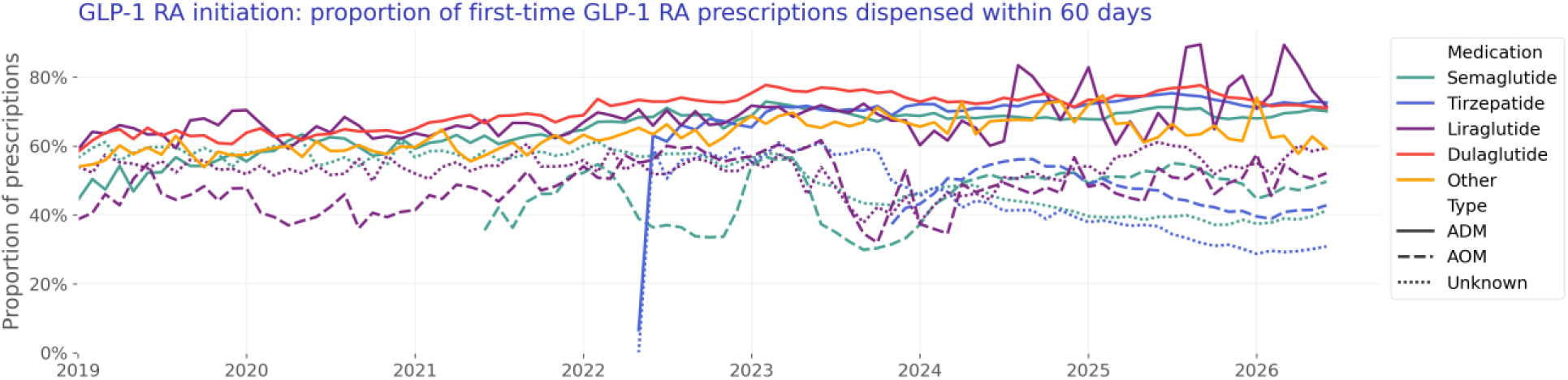

### 60-day initiation over time

We describe the proportion of patients first-prescribed a GLP-1 RA who filled any GLP-1 RA in the subsequent 60 days over time.

Patient characteristics are given in Table 3:

**Table 3:** Patient Characteristics by Dispense Status. Dispense status refers to presence of any GLP-1 RA dispense within 60 days of first GLP-1 RA prescription. Table contains the subset of patients with observable 60-day dispense outcomes.

|  | Dispensed in 60 Days | Not Dispensed in 60 Days | Overall |
| --- | --- | --- | --- |
| N | 2,323,468 (100.00%) | 1,767,397 (100.00%) | 4,090,865 (100.00%) |
| Age Group |  |  |  |
| 18-44 | 544,091 (23.42%) | 528,080 (29.88%) | 1,072,171 (26.21%) |
| 45-64 | 1,131,341 (48.69%) | 810,421 (45.85%) | 1,941,762 (47.47%) |
| 65+ | 648,036 (27.89%) | 428,896 (24.27%) | 1,076,932 (26.33%) |
| Sex |  |  |  |
| Female | 1,452,188 (62.50%) | 1,195,531 (67.64%) | 2,647,719 (64.72%) |
| Male | 871,280 (37.50%) | 571,866 (32.36%) | 1,443,146 (35.28%) |
| Race |  |  |  |
| White | 1,645,407 (70.82%) | 1,302,314 (73.69%) | 2,947,721 (72.06%) |
| Black or African American | 345,111 (14.85%) | 236,357 (13.37%) | 581,468 (14.21%) |
| Asian | 58,246 (2.51%) | 37,155 (2.10%) | 95,401 (2.33%) |
| Unknown | 167,362 (7.20%) | 117,248 (6.63%) | 284,610 (6.96%) |
| Native Hawaiian or Other Pacific Islander | 7,890 (0.34%) | 5,209 (0.29%) | 13,099 (0.32%) |
| American Indian or Alaska Native | 16,556 (0.71%) | 11,753 (0.66%) | 28,309 (0.69%) |
| Other Race | 82,896 (3.57%) | 57,361 (3.25%) | 140,257 (3.43%) |
| Ethnicity |  |  |  |
| Hispanic or Latino | 246,469 (10.61%) | 182,893 (10.35%) | 429,362 (10.50%) |
| Multiethnic | 37 (0.00%) | 22 (0.00%) | 59 (0.00%) |
| Not Hispanic or Latino | 1,735,908 (74.71%) | 1,357,522 (76.81%) | 3,093,430 (75.62%) |
| Unknown | 341,054 (14.68%) | 226,960 (12.84%) | 568,014 (13.88%) |
| Characteristics |  |  |  |
| Type II Diabetes | 1,119,786 (48.19%) | 493,614 (27.93%) | 1,613,400 (39.44%) |
| Overweight or Obesity | 1,519,735 (65.41%) | 1,300,440 (73.58%) | 2,820,175 (68.94%) |
| Medication |  |  |  |
| Semaglutide | 1,181,927 (50.87%) | 945,950 (53.52%) | 2,127,877 (52.02%) |
| Tirzepatide | 701,944 (30.21%) | 602,718 (34.10%) | 1,304,662 (31.89%) |
| Liraglutide | 101,220 (4.36%) | 75,247 (4.26%) | 176,467 (4.31%) |
| Dulaglutide | 310,665 (13.37%) | 123,722 (7.00%) | 434,387 (10.62%) |
| Orforglipron | 2,280 (0.10%) | 4,202 (0.24%) | 6,482 (0.16%) |
| Other | 25,432 (1.09%) | 15,558 (0.88%) | 40,990 (1.00%) |
| Labeled Use |  |  |  |
| ADM | 1,317,408 (56.70%) | 567,521 (32.11%) | 1,884,929 (46.08%) |
| AOM | 519,529 (22.36%) | 586,207 (33.17%) | 1,105,736 (27.03%) |
| Unknown | 486,531 (20.94%) | 613,669 (34.72%) | 1,100,200 (26.89%) |
| Year |  |  |  |
| 2019 | 49,438 (2.13%) | 34,683 (1.96%) | 84,121 (2.06%) |
| 2020 | 61,054 (2.63%) | 41,040 (2.32%) | 102,094 (2.50%) |
| 2021 | 119,558 (5.15%) | 73,041 (4.13%) | 192,599 (4.71%) |
| 2022 | 249,767 (10.75%) | 135,596 (7.67%) | 385,363 (9.42%) |
| 2023 | 432,641 (18.62%) | 276,041 (15.62%) | 708,682 (17.32%) |
| 2024 | 483,832 (20.82%) | 357,949 (20.25%) | 841,781 (20.58%) |
| 2025 | 531,383 (22.87%) | 460,650 (26.06%) | 992,033 (24.25%) |
| 2026 | 395,795 (17.03%) | 388,397 (21.98%) | 784,192 (19.17%) |
| Comorbidity |  |  |  |
| Asthma | 308,974 (13.30%) | 261,516 (14.80%) | 570,490 (13.95%) |
| CKD | 334,029 (14.38%) | 197,797 (11.19%) | 531,826 (13.00%) |
| COPD | 128,477 (5.53%) | 84,026 (4.75%) | 212,503 (5.19%) |
| Hyperlipidemia | 1,169,879 (50.35%) | 872,511 (49.37%) | 2,042,390 (49.93%) |
| Hypertension | 1,214,905 (52.29%) | 884,102 (50.02%) | 2,099,007 (51.31%) |
| Major Depression | 246,000 (10.59%) | 231,546 (13.10%) | 477,546 (11.67%) |

### By Age

As a proxy for Medicare insurance, we segment initiation rate by populations 65+ versus 18-64.

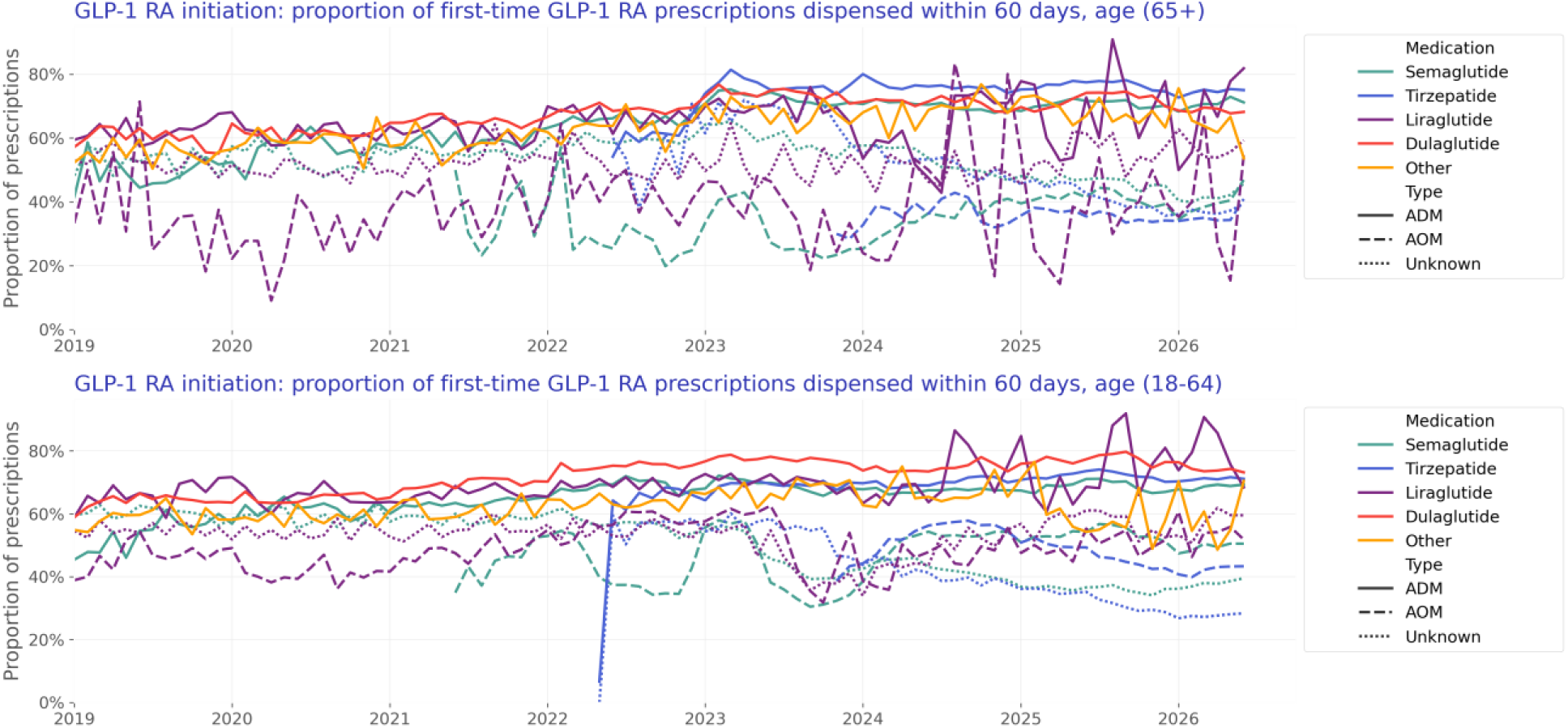

Patient characteristics for patients aged 65+ are given in Table 4:

**Table 4:** Patient Characteristics by Dispense Status, Age 65+. Dispense status refers to presence of any GLP-1 RA dispense within 60 days of first GLP-1 RA prescription. Table contains the subset of patients with observable 60-day dispense outcomes.

|  | Dispensed in 60 Days | Not Dispensed in 60 Days | Overall |
| --- | --- | --- | --- |
| N | 648,036 (100.00%) | 428,896 (100.00%) | 1,076,932 (100.00%) |
| Age Group |  |  |  |
| 65+ | 648,036 (100.00%) | 428,896 (100.00%) | 1,076,932 (100.00%) |
| Sex |  |  |  |
| Female | 362,587 (55.95%) | 251,823 (58.71%) | 614,410 (57.05%) |
| Male | 285,449 (44.05%) | 177,073 (41.29%) | 462,522 (42.95%) |
| Race |  |  |  |
| White | 504,897 (77.91%) | 350,118 (81.63%) | 855,015 (79.39%) |
| Black or African American | 67,211 (10.37%) | 39,064 (9.11%) | 106,275 (9.87%) |
| Asian | 12,645 (1.95%) | 5,888 (1.37%) | 18,533 (1.72%) |
| Unknown | 41,733 (6.44%) | 23,202 (5.41%) | 64,935 (6.03%) |
| Native Hawaiian or Other Pacific Islander | 1,259 (0.19%) | 659 (0.15%) | 1,918 (0.18%) |
| American Indian or Alaska Native | 3,330 (0.51%) | 1,919 (0.45%) | 5,249 (0.49%) |
| Other Race | 16,961 (2.62%) | 8,046 (1.88%) | 25,007 (2.32%) |
| Ethnicity |  |  |  |
| Hispanic or Latino | 43,961 (6.78%) | 23,150 (5.40%) | 67,111 (6.23%) |
| Multiethnic | 4 (0.00%) | 1 (0.00%) | 5 (0.00%) |
| Not Hispanic or Latino | 519,142 (80.11%) | 354,846 (82.73%) | 873,988 (81.16%) |
| Unknown | 84,929 (13.11%) | 50,899 (11.87%) | 135,828 (12.61%) |
| Characteristics |  |  |  |
| Type II Diabetes | 401,446 (61.95%) | 188,802 (44.02%) | 590,248 (54.81%) |
| Overweight or Obesity | 401,066 (61.89%) | 292,377 (68.17%) | 693,443 (64.39%) |
| Medication |  |  |  |
| Semaglutide | 332,720 (51.34%) | 230,044 (53.64%) | 562,764 (52.26%) |
| Tirzepatide | 171,260 (26.43%) | 124,549 (29.04%) | 295,809 (27.47%) |
| Liraglutide | 21,857 (3.37%) | 16,161 (3.77%) | 38,018 (3.53%) |
| Dulaglutide | 109,450 (16.89%) | 49,496 (11.54%) | 158,946 (14.76%) |
| Orforglipron | 1,086 (0.17%) | 1,710 (0.40%) | 2,796 (0.26%) |
| Other | 11,663 (1.80%) | 6,936 (1.62%) | 18,599 (1.73%) |
| Labeled Use |  |  |  |
| ADM | 443,997 (68.51%) | 187,220 (43.65%) | 631,217 (58.61%) |
| AOM | 68,070 (10.50%) | 105,921 (24.70%) | 173,991 (16.16%) |
| Unknown | 135,969 (20.98%) | 135,755 (31.65%) | 271,724 (25.23%) |
| Year |  |  |  |
| 2019 | 14,327 (2.21%) | 11,029 (2.57%) | 25,356 (2.35%) |
| 2020 | 18,119 (2.80%) | 12,905 (3.01%) | 31,024 (2.88%) |
| 2021 | 34,783 (5.37%) | 22,468 (5.24%) | 57,251 (5.32%) |
| 2022 | 63,484 (9.80%) | 34,776 (8.11%) | 98,260 (9.12%) |
| 2023 | 113,936 (17.58%) | 54,358 (12.67%) | 168,294 (15.63%) |
| 2024 | 128,113 (19.77%) | 79,170 (18.46%) | 207,283 (19.25%) |
| 2025 | 149,275 (23.03%) | 107,354 (25.03%) | 256,629 (23.83%) |
| 2026 | 125,999 (19.44%) | 106,836 (24.91%) | 232,835 (21.62%) |
| Comorbidity |  |  |  |
| Asthma | 77,531 (11.96%) | 55,369 (12.91%) | 132,900 (12.34%) |
| CKD | 186,079 (28.71%) | 111,736 (26.05%) | 297,815 (27.65%) |
| COPD | 67,952 (10.49%) | 45,171 (10.53%) | 113,123 (10.50%) |
| Hyperlipidemia | 423,909 (65.41%) | 302,018 (70.42%) | 725,927 (67.41%) |
| Hypertension | 442,958 (68.35%) | 306,980 (71.57%) | 749,938 (69.64%) |
| Major Depression | 66,286 (10.23%) | 52,440 (12.23%) | 118,726 (11.02%) |

Patient characteristics for patients aged 18-65 are given in Table 5:

**Table 5:**
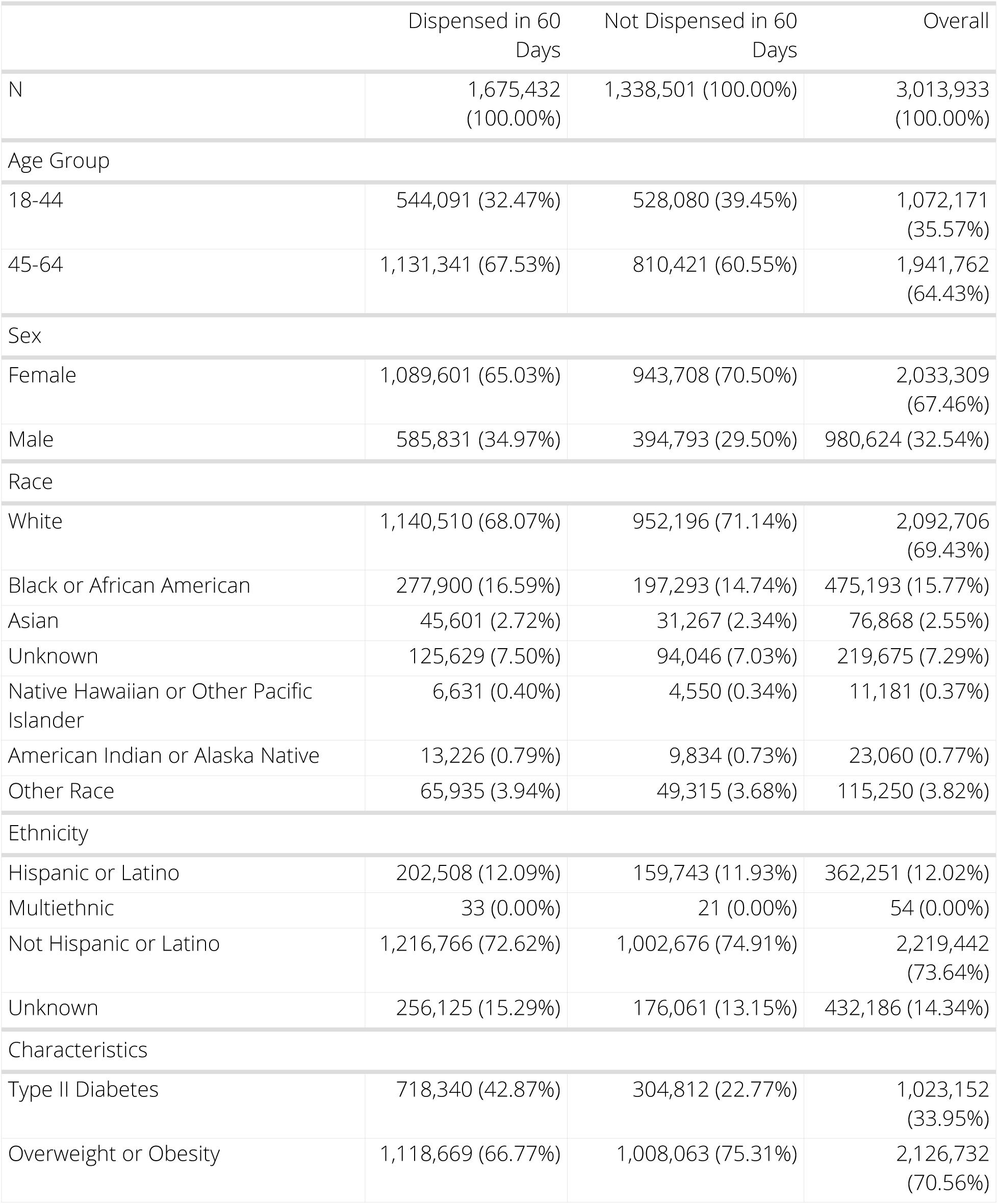
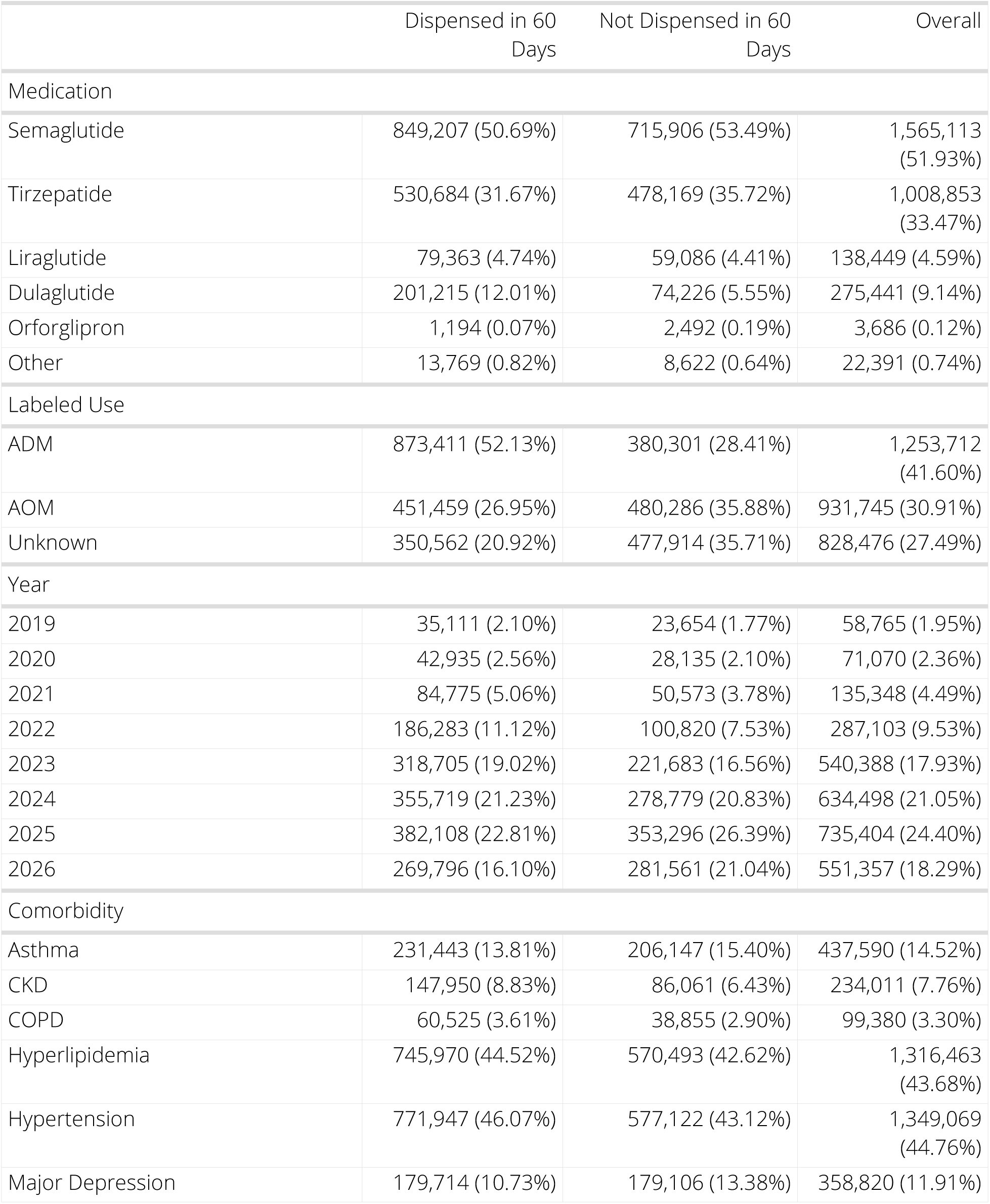
Patient Characteristics by Dispense Status, Age 18-64. Dispense status refers to presence of any GLP-1 RA dispense within 60 days of first GLP-1 RA prescription. Table contains the subset of patients with observable 60-day dispense outcomes.

|  | Dispensed in 60 Days | Not Dispensed in 60 Days | Overall |
| --- | --- | --- | --- |
| N | 1,675,432 (100.00%) | 1,338,501 (100.00%) | 3,013,933 (100.00%) |
| Age Group |  |  |  |
| 18-44 | 544,091 (32.47%) | 528,080 (39.45%) | 1,072,171 (35.57%) |
| 45-64 | 1,131,341 (67.53%) | 810,421 (60.55%) | 1,941,762 (64.43%) |
| Sex |  |  |  |
| Female | 1,089,601 (65.03%) | 943,708 (70.50%) | 2,033,309 (67.46%) |
| Male | 585,831 (34.97%) | 394,793 (29.50%) | 980,624 (32.54%) |
| Race |  |  |  |
| White | 1,140,510 (68.07%) | 952,196 (71.14%) | 2,092,706 (69.43%) |
| Black or African American | 277,900 (16.59%) | 197,293 (14.74%) | 475,193 (15.77%) |
| Asian | 45,601 (2.72%) | 31,267 (2.34%) | 76,868 (2.55%) |
| Unknown | 125,629 (7.50%) | 94,046 (7.03%) | 219,675 (7.29%) |
| Native Hawaiian or Other Pacific Islander | 6,631 (0.40%) | 4,550 (0.34%) | 11,181 (0.37%) |
| American Indian or Alaska Native | 13,226 (0.79%) | 9,834 (0.73%) | 23,060 (0.77%) |
| Other Race | 65,935 (3.94%) | 49,315 (3.68%) | 115,250 (3.82%) |
| Ethnicity |  |  |  |
| Hispanic or Latino | 202,508 (12.09%) | 159,743 (11.93%) | 362,251 (12.02%) |
| Multiethnic | 33 (0.00%) | 21 (0.00%) | 54 (0.00%) |
| Not Hispanic or Latino | 1,216,766 (72.62%) | 1,002,676 (74.91%) | 2,219,442 (73.64%) |
| Unknown | 256,125 (15.29%) | 176,061 (13.15%) | 432,186 (14.34%) |
| Characteristics |  |  |  |
| Type II Diabetes | 718,340 (42.87%) | 304,812 (22.77%) | 1,023,152 (33.95%) |
| Overweight or Obesity | 1,118,669 (66.77%) | 1,008,063 (75.31%) | 2,126,732 (70.56%) |
| Medication |  |  |  |
| Semaglutide | 849,207 (50.69%) | 715,906 (53.49%) | 1,565,113 (51.93%) |
| Tirzepatide | 530,684 (31.67%) | 478,169 (35.72%) | 1,008,853 (33.47%) |
| Liraglutide | 79,363 (4.74%) | 59,086 (4.41%) | 138,449 (4.59%) |
| Dulaglutide | 201,215 (12.01%) | 74,226 (5.55%) | 275,441 (9.14%) |
| Orforglipron | 1,194 (0.07%) | 2,492 (0.19%) | 3,686 (0.12%) |
| Other | 13,769 (0.82%) | 8,622 (0.64%) | 22,391 (0.74%) |
| Labeled Use |  |  |  |
| ADM | 873,411 (52.13%) | 380,301 (28.41%) | 1,253,712 (41.60%) |
| AOM | 451,459 (26.95%) | 480,286 (35.88%) | 931,745 (30.91%) |
| Unknown | 350,562 (20.92%) | 477,914 (35.71%) | 828,476 (27.49%) |
| Year |  |  |  |
| 2019 | 35,111 (2.10%) | 23,654 (1.77%) | 58,765 (1.95%) |
| 2020 | 42,935 (2.56%) | 28,135 (2.10%) | 71,070 (2.36%) |
| 2021 | 84,775 (5.06%) | 50,573 (3.78%) | 135,348 (4.49%) |
| 2022 | 186,283 (11.12%) | 100,820 (7.53%) | 287,103 (9.53%) |
| 2023 | 318,705 (19.02%) | 221,683 (16.56%) | 540,388 (17.93%) |
| 2024 | 355,719 (21.23%) | 278,779 (20.83%) | 634,498 (21.05%) |
| 2025 | 382,108 (22.81%) | 353,296 (26.39%) | 735,404 (24.40%) |
| 2026 | 269,796 (16.10%) | 281,561 (21.04%) | 551,357 (18.29%) |
| Comorbidity |  |  |  |
| Asthma | 231,443 (13.81%) | 206,147 (15.40%) | 437,590 (14.52%) |
| CKD | 147,950 (8.83%) | 86,061 (6.43%) | 234,011 (7.76%) |
| COPD | 60,525 (3.61%) | 38,855 (2.90%) | 99,380 (3.30%) |
| Hyperlipidemia | 745,970 (44.52%) | 570,493 (42.62%) | 1,316,463 (43.68%) |
| Hypertension | 771,947 (46.07%) | 577,122 (43.12%) | 1,349,069 (44.76%) |
| Major Depression | 179,714 (10.73%) | 179,106 (13.38%) | 358,820 (11.91%) |

## Discussion and Conclusion

This analysis summarized GLP-1 RA prescribing trends and patient characteristics from January 2019 to September 2026. During this period 4,458,532 patients were prescribed a total of 23,616,330 total prescriptions. ADMs accounted for 65.7% of prescriptions, and AOMs accounted for 34.3%, among prescriptions where use was known. A larger proportion of ADMs, compared to AOMs, were dispensed within 60 days: 69.9% of first-time ADM prescriptions and 47.0% of first-time AOM prescriptions had a fill within 60 days.

Several limitations of our analysis should be noted. All data are preliminary and may change as additional data are obtained. These findings are consistent with data accessed October 23, 2026. Data presented in this analysis represent raw counts and/or rates, and post-stratification methods have not been conducted. In addition, cohorts with small counts may be suppressed during the de-identification process leading to the appearance of zero patients for a given time period. Given relatively large numbers of patients prescribed and dispensed GLP-1 RA, we do not expect substantial suppression in the most recent periods. The unknowns in this report either indicate the value was not included in the individual’s electronic health record or that it was excluded from the data to protect an individual’s identity as a part of Truveta’s commitment to privacy (Truveta, 2024). Finally, this report is a description of prescribing trends and patient characteristics only. Results should not be interpreted as causal.

## Data Availability

The data used in this study are available to all Truveta subscribers and may be accessed at studio.truveta.com.

## Acknowledgements

The authors thank Dr. Ty Gluckman for his contributions and feedback on describing GLP-1 RA prescribing trends.

